# Association of homelessness with COVID-19 positivity among individuals visiting a testing center

**DOI:** 10.1101/2021.01.04.20248661

**Authors:** Tara Kiran, Amy Craig-Neil, Paul Das, Joel Lockwood, Ri Wang, Nikki Nathanielsz, Esther Rosenthal, Stephen W. Hwang

## Abstract

We conducted a chart audit of all patients attending an inner-city COVID-19 testing centre in Toronto, Canada between March and April 2020. Of the 2050 unique individuals tested, 214 (10.4%) were homeless. People experiencing homelessness were more likely to test positive for COVID-19 compared to those not experiencing homelessness even after adjustment for age, sex, and the presence of any medical co-morbidity (15.4% vs. 6.7%, p<0.001; OR 2.41, 95% CI 1.51 to 3.76, p<0.001).

## Introduction

People experiencing homelessness are at higher risk of acquiring SARS CoV-2 as lack of safe housing makes it difficult to practice physical distancing, hand hygiene, and other preventive measures.^1^ People experiencing homelessness also have higher rates of chronic conditions, making them more vulnerable to COVID-19 complications.^2^ Early in the pandemic, some regions began conducting mobile outreach testing in shelters and detected high rates of infection among asymptomatic residents, especially when there was a known positive case in the shelter.^3,4^ However, it is unclear how often people experiencing homelessness are visiting testing centres and how their test positivity rate differs from that of others visiting the same centre.

The St. Michael’s Hospital COVID-19 Assessment Centre (CAC) was one of 116 testing centres opened in Ontario, Canada, shortly after the pandemic began. It is located in Toronto’s urban core where a large proportion of the city’s homeless population resides. This study examines the association between homelessness and test positivity among people seen at the assessment center.

## Methods

We conducted a retrospective chart audit of all patients tested for COVID-19 at the St. Michael’s Hospital CAC from its opening on March 16, 2020 until April 30, 2020. Testing was free for all individuals. Testing criteria changed according to provincial government direction^5^ and was largely limited to symptomatic people who were at high-risk of acquiring COVID-19 due to vulnerable residence, occupation, or high-risk exposure. Vulnerable residence included those unhoused or in homeless shelters. In mid-April, asymptomatic individuals began being tested in specific circumstances (e.g. local outbreak, clinical exposure). We did not include results from the CAC’s outreach testing done at shelters.

Data were collected on a standardized form by registered nurses, nurse practitioners or physicians. We classified people as homeless based on data from the chart and the hospital registration address field. We used a Chi-squared test or Mann-Whitney test to compare demographics, medical co-morbidities, symptoms, and test positivity between people who did and did not experience homelessness. We performed a logistic regression analysis to estimate the odds of testing positive for COVID-19 for people who were and were not homeless after adjustment for age, sex, and medical co-morbidity.

## Results

Between March and April 2020, 214 (10.4%) of 2050 unique individuals who were tested at the St. Michael’s Hospital CAC were homeless. People experiencing homelessness were more likely to be male (75.7% vs 37.0%, p<0.001) and less likely to have a health insurance card (71.5% vs. 97.6%, p<0.001) (Table 1). There was no statistical difference in mean age, but the age distribution was different (p<0.001) with fewer people experiencing homelessness between age 25 to 49. There were no statistical differences in reported symptoms but people experiencing homelessness were more likely to have at least one medical co-morbidity (70.3% vs. 53.4%, p<0.001) and abnormal vital sign (38.1% v. 26.0%, p<0.01) compared to those not experiencing homelessness.

**Table 1.** Comparison of demographic characteristics, symptoms, medical co-morbidity, and vital signs between people who did and did not experience homelessness

|  | Homeless<br>(n=214) | Not homeless<br>(n=1836) | All<br>(n=2050) | P-value |
| --- | --- | --- | --- | --- |
| Age, Median (IQR) | 40.3 (31.0-55.5) | 41.7 (32.1-54.0) | 41.5 (32.1-54.1) | 0.64 |
| Age category |  |  |  | <0.001 |
| 0-15 | 4 (1.9%) | 14 (0.8%) | 18 (0.9%) |  |
| 16-24 | 23 (10.8%) | 90 (4.9%) | 113 (5.5%) |  |
| 25-49 | 111 (51.9%) | 1130 (61.6%) | 1241 (60.5%) |  |
| 50-64 | 58 (27.1%) | 497 (27.1%) | 555 (27.1%) |  |
| 65+ | 18 (8.4%) | 105 (5.7%) | 123 (6.0%) |  |
| Sex |  |  |  | <0.001 |
| Female | 52 (24.3%) | 1155 (63.0%) | 1207 (59.0%) |  |
| Male | 162 (75.7%) | 678 (37.0%) | 840 (41.0%) |  |
| OHIP available | 153 (71.5%) | 1792 (97.6%) | 1945 (94.9%) | <0.001 |
| Symptoms |  |  |  |  |
| Any symptoms | 172 (83.1%) | 1563 (85.8%) | 1735 (85.6%) | 0.34 |
| No symptoms | 35 (16.9%) | 258 (14.2%) | 293 (14.5%) |  |
| Cough | 100 (48.3%) | 892 (49.0%) | 992 (48.9%) | 0.85 |
| Fever | 27 (13.0%) | 193 (10.5%) | 220 (10.8%) | 0.26 |
| Shortness of Breath | 25 (12.1%) | 229 (12.6%) | 254 (12.5%) | 1.00 |
| Other | 91 (44.0%) | 827 (45.4%) | 918 (45.3%) | 1.00 |
| Medical co-morbidity |  |  |  |  |
| Any co-morbidity | 135 (70.3%) | 911 (53.4%) | 1046 (55.1%) | <0.001 |
| No co-morbidity | 57 (29.7%) | 796 (46.6%) | 853 (44.9%) |  |
| Chronic lung disease | 25 (13.0%) | 179 (10.5%) | 204 (10.7%) | 0.85 |
| Diabetes | 14 (7.3%) | 133 (7.8%) | 147 (7.7%) | 0.24 |
| Heart disease or stroke | 14 (7.3%) | 83 (4.9%) | 97 (5.1%) | 0.76 |
| Immunosuppressed | 9 (4.7%) | 61 (3.6%) | 70 (3.7%) | 1.00 |
| Smoker | 85 (44.3%) | 190 (11.1%) | 275 (14.5%) | <0.001 |
| Other | 66 (34.4%) | 485 (28.4%) | 551 (29.0%) | 0.39 |
| Any abnormal vital sign* | 48 (38.1%) | 288 (26.0%) | 336 (27.2%) | <0.01 |

\*Abnormal vital sign defined as heart rate>110, oxygen saturation<92%, and/or respiratory rate>24

People experiencing homelessness were more likely to test positive for COVID-19 compared to those not experiencing homelessness (15.4% vs. 6.7%, p<0.001). People experiencing homelessness had higher odds for testing positive even after adjustment for age, sex, and the presence of any medical co-morbidity (OR 2.41, 95% CI 1.51 to 3.76, p<0.001) (Table 2).

**Table 2.**
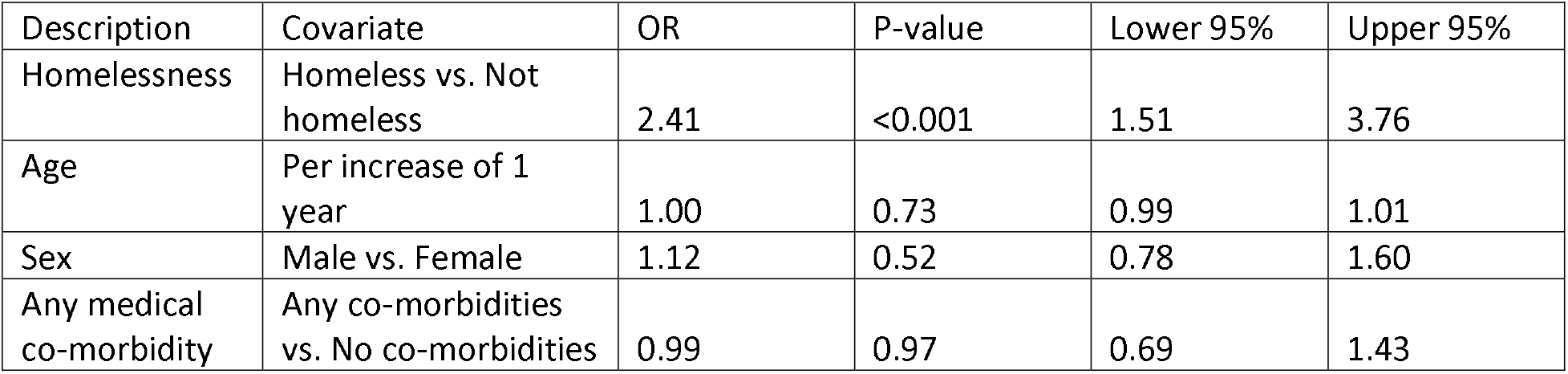
Odds of individuals experiencing homelessness testing positive for COVID-19 compared to individuals not experiencing homelessness after adjustment for age, sex, and any medical co-morbidity

| Description | Covariate | OR | P-value | Lower 95% | Upper 95% |
| --- | --- | --- | --- | --- | --- |
| Homelessness | Homeless vs. Not homeless | 2.41 | <0.001 | 1.51 | 3.76 |
| Age | Per increase of 1 year | 1.00 | 0.73 | 0.99 | 1.01 |
| Sex | Male vs. Female | 1.12 | 0.52 | 0.78 | 1.60 |
| Any medical co-morbidity | Any co-morbidities vs. No co-morbidities | 0.99 | 0.97 | 0.69 | 1.43 |

## Discussion

In this study of individuals visiting a COVID-19 testing centre early in the pandemic, people experiencing homelessness had more than twice the odds of testing positive than those not experiencing homelessness. The higher positivity was present even when accounting for differences in age, sex, and medical co-morbidity. A limitation of this study is that it reflects testing at a single centre early in the pandemic when testing was largely limited to those living or working in high-risk settings. The City of Toronto has since moved more than 3500 individuals experiencing homelessness into spaces that allow for physical distancing.^6^ Research is needed to understand whether these efforts have lowered the rates of COVID-19 infection among people who are unhoused. Our results confirm that people experiencing homelessness are at high-risk of COVID-19 and that targeted efforts are needed to reduce transmission rates.

## Data Availability

Due to the nature of this research, participants of this study did not agree for their data to be shared publicly, so supporting data is not available.

## Acknowledgements

We are grateful to Linh Luong and Tadios Tibebu who spent many hours extracting data from paper charts. We thank the many individuals involved in supporting the mobile outreach testing, particularly Dana Whitham, Nicole Gichuru, Chantel Marshall and Linda Jackson, whose leadership and guidance made the outreach possible. We would like to especially acknowledge the daily hard work by staff at the COVID-19 Assessment Centre at St. Michael’s Hospital, Sherbourne Health, and the shelter sites.

## References

1. Perri M, Dosani N, Hwang SW. Covid-19 and people experiencing homelessness: Challenges and mitigation strategies. CMAJ. 2020;192(26):E716–E719.

2. Fazel S, Geddes JR, Kushel M. The health of homeless people in high-income countries: Descriptive epidemiology, health consequences, and clinical and policy recommendations. The Lancet. 2014;384(9953):1529–1540.

3. Baggett TP, Keyes H, Sporn N, Gaeta JM. Prevalence of sars-cov-2 infection in residents of a large homeless shelter in boston. Jama. 2020;323(21):2191–2192.

4. Mosites E, Parker EM, Clarke KE, et al. Assessment of sars-cov-2 infection prevalence in homeless shelters—four us cities, march 27–april 15, 2020. Morbidity and Mortality Weekly Report. 2020;69(17):521.

5. Ministry of Health and Long-Term Care. Covid-19 guidance for the health sector. http://www.health.gov.on.ca/en/pro/programs/publichealth/coronavirus/2019_guidan ce.aspx#health. Accessed December 1, 2020.

6. City of toronto covid-19 response for people experiencing homelessness. Aug 7, 2020; https://www.toronto.ca/news/city-of-toronto-covid-19-response-for-people-experiencing-homelessness/. Accessed Sep 14, 2020.

